# Exploring the Impact of Aphasia Severity on Employment, Social Participation, and Quality of Life

**DOI:** 10.1101/2025.01.08.25320231

**Authors:** Mika Konishi, Michitaka Funayama, Fumie Saito, Yoshitaka Nakagawa, Naomi Fujinaga, Masayo Urano, Masanori Osumi, Shu Harayama, Masako Tateishi, Jun Tanemura, Masaru Mimura

**Affiliations:** Department of Neuropsychiatry, Keio University School of Medicine, Tokyo, Japan; Department of Neuropsychiatry, Keio University School of Medicine, Tokyo, Japan Department of Neuropsychiatry, Ashikaga Red Cross Hospital, Tochigi, Japan; Department of Rehabilitation, Edogawa Hospital, Tokyo, Japan; Department of Rehabilitation, Tokyo Metropolitan Rehabilitation Hospital, Tokyo, Japan; School of Health Sciences, Tokyo University of Technology, Tokyo, Japan; Department of Rehabilitation, Kasumigaseki-minami Hospital, Saitama, Japan; Department of Speech-Language pathology and Audiology, Faculty of Rehabilitation, Kawasaki University of Medical Welfare, Okayama, Japan; Japanese Association of Speech-Language-Hearing Therapists, Tokyo, Japan; Biwako Professional University of Rehabilitation, Shiga, Japan; Center for Preventive Medicine, Keio University, Tokyo, Japan

**Keywords:** aphasia severity, employment, social participation, quality of life, social environmental barriers

## Abstract

**Background:** The primary challenge faced by patients with aphasia is their difficulties in communicating, which likely contributes to lower employment rates, decreased social participation, and a decline in quality of life. Surprisingly, few studies have investigated the relationship between aphasia severity and these outcomes, particularly employment status. In this study, we addressed this gap by examining these socio-occupational outcomes as well as quality of life in individuals with chronic aphasia.

**Methods:** A cohort of 136 individuals with chronic aphasia following cerebrovascular diseases was recruited and investigated in a cross-sectional study. A multiple logistic regression model was used for employment status, and multiple linear regression models were used for both social participation levels and quality of life. Explanatory variables included both individual’s functions, such as aphasia severity, non-linguistic cognitive function, apathy levels, and mobility, as well as levels of social environmental barriers.

**Results:** Aphasia severity had a significant negative impact on two outcomes: employment status and quality of life. Additionally, social environmental barriers negatively affected quality of life. Lower mobility, male gender, and older age were related to reduced social participation levels. Employment status was not related to social participation levels nor quality of life.

**Conclusion:** This study found that the severity of aphasia significantly impacts individuals’ occupational engagement, as well as their quality of life. Our findings shed light on potential treatment options during acute phases of stroke, linguistic rehabilitation, and occupational support for individuals with aphasia.

## Introduction

Individuals with aphasia encounter numerous challenges and limitations in their daily lives due to impaired communication abilities. As a result, they often struggle to maintain employment status and participate socially^1^, which affects their economic, vocational, and social lives, as well as their overall quality of life^2^. These difficulties are presumed to worsen with increasing aphasia severity^2^. Therefore, understanding the severity of aphasia is crucial for comprehending its impact on socio-occupational outcomes, more so than merely noting the presence or absence of aphasia. However, studies specifically focusing on aphasia severity in relation to socio-occupational engagement, especially employment status, are scarce. Understanding the impact of aphasia severity is particularly important given the recent development of intensive treatments for acute ischemic stroke^3, 4^, the leading cause of aphasia. These treatments include acute reperfusion therapies such as intravenous thrombolysis, endovascular thrombectomy, and combined therapy (intravenous thrombolysis with endovascular thrombectomy) ^3, 4^. As these treatments can now reduce aphasia severity^5, 6^, comprehending its impact on socio-occupational status during the chronic stage can aid in treatment decisions, especially for individuals whose occupations might be affected by aphasia severity. Understanding the impact of aphasia severity on socio-occupational engagement is also crucial for determining specific methods in linguistic rehabilitation and occupational support for individuals with aphasia^1, 7^.

Most previous research on employment status after cerebrovascular disease did not specifically focus on aphasia or its severity, but rather categorized its presence as one of several cognitive dysfunctions^8–12^. These studies indicated that the presence of aphasia negatively impacts employment status along with other dysfunctions, such as age, education levels, depression, impaired mobility, non-linguistic cognitive dysfunctions, and social environmental barriers. Naess et al. ^13^ focused on the presence of aphasia and found that, among stroke survivors, individuals with aphasia were less employed than those without aphasia. Surveys on employment status among individuals with aphasia^1, 14^ also underscore significant challenges faced by individuals with aphasia. While these past reports and surveys emphasize the presence of aphasia as a crucial factor for poor employment status, they do not delve into the specific impact of aphasia severity on employment.

In terms of social participation, Dalemans et al. ^15^ found a correlation between aphasia severity and social participation, alongside age, gender, and functional abilities. This study appears to be the sole investigation focusing explicitly on aphasia severity in this context. Lee et al. ^16^ and Ohata et al. ^17^ demonstrated that while they did not specifically consider the severity of aphasia, the presence of aphasia negatively affects social participation levels compared to healthy individuals.

Compared to studies on employment status and social participation levels, research on the relationship between the severity of aphasia and quality of life among patients with aphasia has been more extensive. Studies have identified several factors associated with quality of life in individuals with aphasia, including the severity of aphasia, depression, gender, age, education levels, social environmental barriers, functional abilities, and fatigue ^1, 16, 18–20^.

This study aims to address gaps in the literature, particularly focusing on the relationship between the severity of aphasia and employment status. To this end, we conducted a comprehensive cross-sectional study to examine how the severity of aphasia impacts employment status, social participation, and associated quality of life among individuals with aphasia. Additionally, we explored the influence of employment status on social participation and quality of life in this patient group.

## Methods

### Participants

The ethical aspects of this study were reviewed and approved by the Human Research Ethics Committee at Keio University School of Medicine (2021-1163-5). Informed consent was obtained from all participants before the study commenced. Participants were recruited between June 2019 and March 2024 from neuropsychiatry clinic affiliated with Keio University hospital, 25 rehabilitation facilities, and two cognitive dysfunction clinics affiliated with general hospitals across Japan. The participants in this study were individuals who had experienced cerebrovascular diseases and met the following criteria: they were native Japanese individuals aged over 20 years, diagnosed with aphasia by both a medical doctor and a speech-language pathologist (each with more than 5 years of experience in aphasia diagnosis and/or treatment), more than 6 months post-stroke, medically stable, capable of independent self-care (including eating, grooming, bathing, dressing, and toileting), had no known history of neurological or psychiatric disease prior to the cerebrovascular diseases, and had no neurological comorbidities after the cerebrovascular diseases, such as neurodegenerative diseases. To include individuals with severe aphasia, primary caregivers were required to participate and respond to questionnaires on behalf of aphasic participants. This was particularly necessary for the Craig Hospital Inventory of Environmental Factors (CHIEF) ^21^, which assesses social environmental barriers affecting individuals with disabilities. These questionnaires can be challenging for individuals with aphasia because they were not specifically designed for individuals with cognitive dysfunction. Conversely, the assessments of social participation and quality of life were conducted directly with individuals with aphasia^1, 16, 18–20^. These assessments, which will be described below, were specifically designed for individuals with cognitive dysfunction, including those with aphasia, making them relatively easier to understand and answer.

### Assessments

We included both personal and socio-environmental explanatory variables to account for employment status, social participation, and quality of life among individuals with aphasia following cerebrovascular diseases. The personal explanatory variables involve the abilities of individuals with aphasia, which encompass aphasia severity, neuropsychological, psychiatric, and physical functions. The socio-environmental variables include social environmental barriers, which affect the individuals. We also considered demographic information. These variables were examined in previous studies on socio-occupational outcomes and quality of life in individuals with aphasia ^1, 8–12, 15, 18–20, 22^.

### Outcomes

#### Employment Status

This assessment was given to working-age individuals under 65 years old who are at least 18 months past the onset of aphasia. This period aligns with the expiration of Sickness and Injury Benefits in Japan, a health insurance provision for extended paid leave due to injury or illness, marking the maximum allowable period of employment despite the inability to work (https://www.mhlw.go.jp/content/12401000/000619554.pdf). Additionally, this timeframe is consistent with that used in previous surveys on employment status in individuals with chronic aphasia^1, 22^, which recruited individuals who were at least 13 months post-onset. We categorized employment status into two scales: ‘employed,’ which includes both full-time and part-time workers, and ‘unemployed,’ which also covers those undergoing vocational training and employment under disability schemes, both of which are operated with subsidies. We chose not to include the Productivity subscale from the Community Integration Questionnaire^15, 23^ , which was used for Productivity among individuals with aphasia in previous studies^15, 23^. This decision was based on the fact that the Productivity subscale encompasses factors beyond employment status, including volunteer activities, training, and school attendance, which are not the primary focus of our manuscript. Instead, as described below, we used only the Social Integration subscale from the Community Integration Questionnaire to assess social participation levels.

#### Social Participation

We assessed levels of social participation in individuals who are at least 6 months past the onset of aphasia. This timeframe aligns with a previous study^16^ and corresponds to the typical discharge limit for patients with cerebrovascular disease from the recovery phase rehabilitation ward in Japan (https://www.mhlw.go.jp/content/12404000/000864213.pdf). We utilized the Japanese version of the Social Integration subscale from the Community Integration Questionnaire^24^, which was originally developed for individuals with traumatic brain injuries and later adapted for those with aphasia^15, 23^. The Social Integration subscale covers six items: personal finance management, shopping, leisure activities outside the home, visiting friends or relatives, engaging in leisure activities with others, and having a confidant. Each item is scored on a scale of 0 to 2, with a maximum total score of 12. While the Community Integration Questionnaire also includes Home Activities and Productivity subscales, we focused specifically on the Social Integration subscale in this manuscript to align with our research objectives regarding social participation among individuals with aphasia. Additionally, this Social Integration subscale, unlike the others, has been reported to be closely linked to the quality of life among individuals with aphasia in one study^16^.

#### Quality of Life

We assessed the quality of life for individuals who are at least 6 months past the onset of aphasia using the Japanese version of the Stroke and Aphasia QOL Scale-39 (SAQOL-39) ^25, 26^, a validated and reliable tool for individuals with aphasia worldwide and in Japan. The SAQOL-39 comprises 39 questions distributed across four domains: physical (17 items), psychosocial (11 items), communication (7 items), and energy (4 items). Each item is evaluated on a five-point scale from 1 to 5. The average score is calculated by dividing the total score by 39, the total number of items on this questionnaire. We also performed a sub-assessment of the communication domain using the 7 items related to communication, as aphasia severity predominantly affects communication abilities. The communication domain score was calculated by dividing the total score of these 7 items by 7.

### Explanatory Variables

#### Aphasia Severity

The severity of aphasia was determined using the Standard Language Test of Aphasia (SLTA), a widely-used and validated assessment tool for aphasia in Japan for over half a century^27, 28^. The SLTA assesses aphasia severity on a scale of 0 to 10^27, 28^ based on Guttman’s scalogram analysis or cumulative scaling^29^, which is a deterministic model. The SLTA assesses three linguistic components: comprehension, speech, and reading and writing of written language. Each linguistic component of the SLTA comprises 3 to 4 subcategories, ranked from least to most difficult, resulting in a total of 10 subcategories. For instance, in the comprehension component, if a patient achieved an 80% passing mark in easy word comprehension, they were assigned one grade for a subcategory requiring the simplest comprehension level. Similarly, if a patient attained an 80% passing mark in sentence comprehension, they were assigned one grade for a subcategory demanding the highest level of comprehension. These 10 subcategories are then converted into a total score of 10 grades to assess aphasia severity, with a grade of 0 indicating the poorest performance and a grade of 10 reflecting the best^27, 28^.

#### Non-Linguistic Cognitive Function

The Raven’s Colored Progressive Matrices test, a non-verbal assessment, was used to evaluate non-linguistic cognitive function. The results from this test often correlate well with those from verbal intelligence tests, making it a commonly used tool for assessing non-linguistic cognitive function in individuals with aphasia^30^.

#### Physical and Psychosocial factors and Demographics

We assessed physical function (mobility) using the Japanese version of the Walk/Wheelchair subscale of the Functional Independent Measure (FIM) ^31, 32^ scored on a scale from 1 (worst) to 7 (best).

#### Social Environmental Barriers

We utilized the Craig Hospital Inventory of Environmental Factors (CHIEF), a questionnaire designed to assess inhibitory environmental factors affecting individuals with disabilities^17, 21^. The CHIEF focuses on quantifying barriers experienced by people with disabilities, encompassing 25 items distributed across five domains of social environmental factors: Policies, Physical and Structural, Work and School, Attitudes and Support, and Services and Assistance. Each CHIEF item is scored based on the product of a frequency score (ranging from never = 0 to daily = 4) and a magnitude of impact score (ranging from no problem = 0 to big problem = 2), resulting in an item score ranging from 0 to 8. Higher scores indicate a greater frequency and/or magnitude of environmental barriers, with a maximum score of 200. The CHIEF is also recognized as a valuable scale for evaluating environmental factors in individuals with aphasia^33^. We did not use the sub-assessment within the CHIEF, i.e., the Work and School domain, as an explanatory variable specifically for employment status. This is because employment difficulties in individuals with aphasia arise not only from items in the Work and School domain but also from items in other domains, such as information access and digital literacy.

#### The Other Variables

Psychiatric factors included presence of depression (determined by antidepressant medication at the time of investigation) and levels of apathy. Depression has sometimes been associated with quality of life among individuals with aphasia^16, 19^. Apathy levels were included in our analysis because this assessment has been utilized and found to be closely associated with poor rehabilitation outcomes after cerebrovascular diseases^34^ and low employment status among individuals with traumatic brain injury^35^. Apathy levels were assessed using the Clinical Assessment of Spontaneity, a validated and reliable tool in Japan^36^, specifically the Clinical Overall Spontaneity Assessment, ranging from 0 (no apathy) to 5 (extremely severe apathy). We did not include levels of fatigue as an explanatory variable, which are closely related to apathy levels^37^ and were found to be associated with social participation in individuals with aphasia^38^. This decision was made to avoid circularity, as fatigue levels were already encompassed within the energy domain of the SAQOL, a measure of quality of life that is among our outcome measures. Demographic factors investigated were age, months post-onset, gender, marital status (single or married), and education level (in years). Marital status was included because it is often investigated in socio-occupational and quality of life studies on individuals with aphasia^12, 20^.

### Statistical Analysis

Initially, we examined the relationship between each explanatory variable and each outcome. For employment status, we compared employed and non-employed individuals using the Chi-square test or Fisher’s exact test for categorical variables. The Chi-square test was used when expected cell counts were > 5, while the Fisher’s exact test was used when expected cell counts were ≦ 5. For numerical variables, we used the Mann-Whitney U test, since these data did not follow a normal distribution. Regarding social participation levels and quality of life, we explored the relation between each explanatory variable and each outcome using simple linear regression analysis for numerical variables and Mann-Whitney U test for categorical variables. Despite employment status and social participation being the primary outcomes, we also investigated their relationship with quality of life, as social participation is known to influence quality of life among individuals with aphasia^16^, and employment status might have a similar effect. Also, we investigated the relation between employment status and social participation levels.

Secondly, to control for confounding factors, we utilized the multiple logistic regression analysis for employment status and the multiple linear regression analysis for social participation levels and quality of life. We included only those variables with low p-values from the previously described individual comparison analyses, specifically those with *p* < 0.10. This decision was made after considering the small number of individuals assessed for employment status—those who were under 65 years old and more than 18 months post-onset—the number of explanatory variables was generally limited to less than one-tenth of the number of participants^39^. To specifically investigate the communication aspect of quality of life, the SAQOL-39 was assessed using both the overall total score and the subscale score of the communication domain. Statistical analyses were performed using SPSS version 29.0.1.0 and R 4.1.1, and a P-value of < 0.05 was considered statistically significant.

## Results

Table 1 presents the socio-demographic, neuropsychological, psychiatric, and physical data for two cohorts: one for employment status (≧18months post-onset) and the other for social participation and quality of life (≧6 months post-onset). The participants were predominantly male (>70%), with an average age of approximately 60, and generally had good mobility. The primary etiology was cerebral infarction, affecting approximately 60% of the participants, followed by cerebral hemorrhage and subarachnoid hemorrhage. Aphasia severity averaged 7.7 out of 10, indicating mild to moderate aphasia, but with a wide standard deviation. Non-linguistic cognitive function was relatively preserved, with an average score of approximately 30 out of 36, compared to the normal range of 29.2±5.4 for healthy individuals in their 60s. This measure also had a wide standard deviation. Apathy was observed in roughly 20% of the participants, while depression was noted in fewer than 5%.

**Table 1.** Outcomes and Explanatory Variables of the Participants.

|  | **Cohort for employment status**<br>**(working age participants**<br>**with >18 months post-onset,**<br>**N=56)** | **Cohort for social participation**<br>**levels and quality of life ( > 6**<br>**months post-onset, N=136)** |
| --- | --- | --- |
| **Age (years)** | 53.1±10.0 | 60.6±13.5 |
| **Male gender** | 40 (71%) | 102 (75%) |
| **Education (years)** | 13.9±2.2 | 13.9±2.3 |
| **Marital status (% for**<br>**married)** | 56% | 65% |
| **Post onset (months)** | 92.7±89.9 | 67.1±81.1 |
| **Etiology** | Stroke 57%<br>Hemorrhage 30%<br>SAH 13% | Stroke 63%<br>Hemorrhage 27%<br>SAH 10% |
| **Mobility (FIM)**<br>**(0, worst; 7, best)** | 6.7±0.7 | 6.8±0.6 |
| **Apathy level**<br>**(0, best; 5, worst)** | 0.39±0.66 | 0.34±0.59 |
| **Depression** | 2 (3.6%) | 5 (3.7%) |
| **Severity of Aphasia**<br>**(SLTA) (0, worst; 10, best)** | 7.1 ± 2.6 | 7.4 ± 2.5 |
| **Non-Verbal Intelligence**<br>**(RCPM) (0, worst; 36, best)** | 30.3 ± 5.2 | 29.3 ± 6.4 |
| **Employment Status** | 13 (23%)<br>(regular 11; part-time 2) | NA |
| **Social Participation**<br>**(CIQ) (0, lowest; 12, highest)** | 5.1 ± 2.7 | 5.0 ± 2.5 |
| **Quality of Life**<br>**(SAQOL) (0, lowest; 5, highest)** | 3.6 ± 0.6 | 3.7 ± 0.6 |
| **Communication domain in**<br>**Quality of Life**<br>**(SAQOL communication)**<br>**(0, lowest; 5, highest)** | 2.9 ± 0.9 | 3.1 ± 1.0 |
| **Social Environmental**<br>**Barriers**<br>**(CHIEF)**<br>**(0, lowest; 200 highest)** | 30.3 ± 26.4 | 19.0 ± 24.3 |

Note: CHIEF; Craig Hospital Inventory of Environmental Factors, CIQ; Community Integration Questionnaire, FIM; Functional Independence Measure, NA; not applicable, RCPM; Raven's Coloured Progressive Matrices, SAH; subdural hemorrhage, SAQOL; Stroke and Aphasia QOL Scale-39, SLTA; Standard Language Test of Aphasia,

**Table 2.**
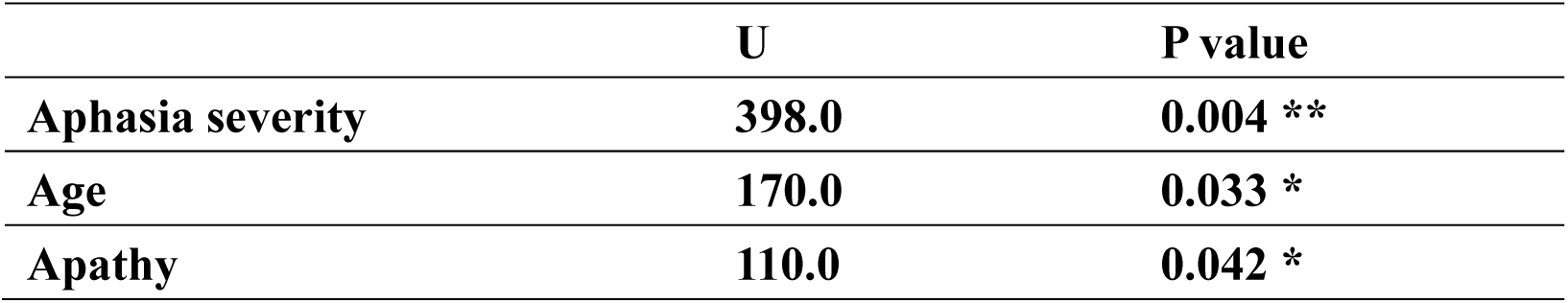
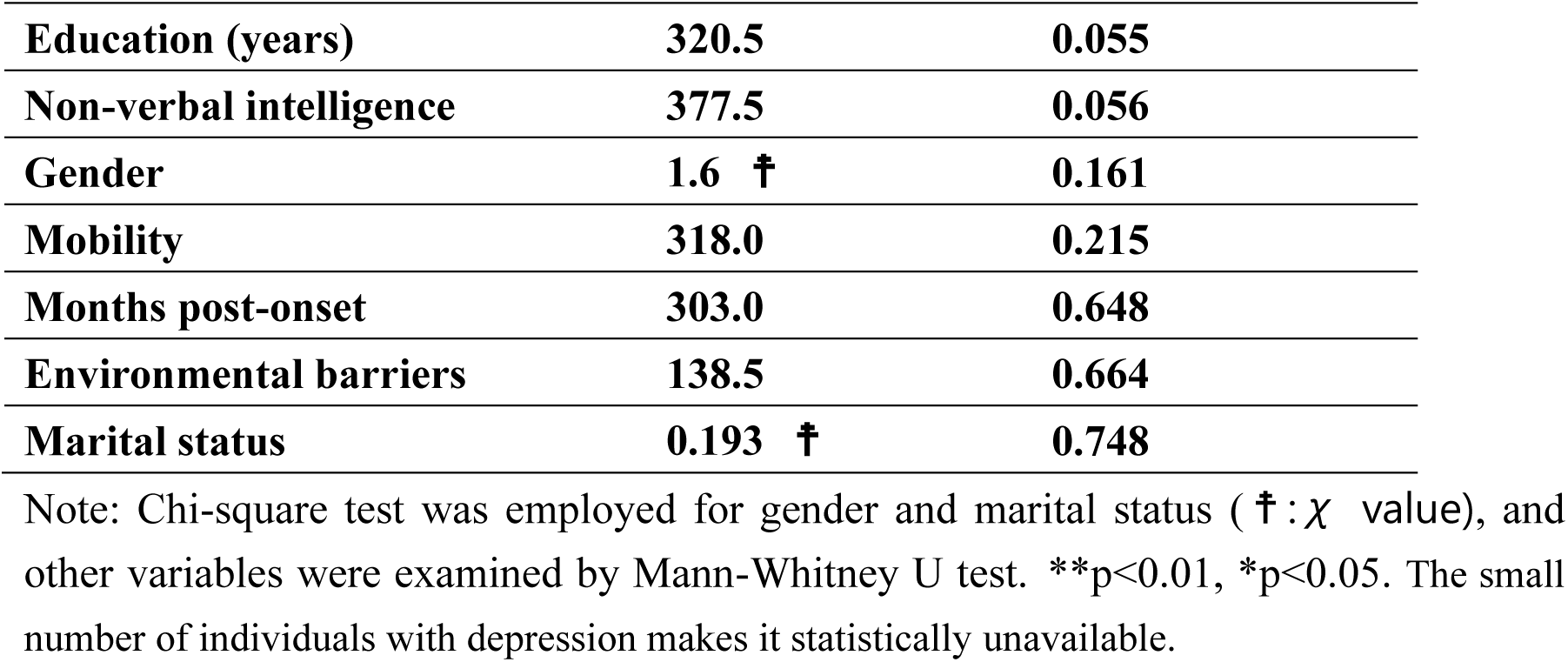
Relationship between employment status and explanatory variables.

**Table 3.** Correlations between Explanatory Variables and Social Participation, Quality and Life, or Communication Domain in Quality of Life.

|  | Social Participation |  | Quality of Life |  | Communication |
| --- | --- | --- | --- | --- | --- |
|  | r | P value | r | P value | r |
| **Aphasia Severity** | **0.145** | **0.140** | **0.341** | **< 0.001\*\*** | **0.204** |
| **Age** | **-0.227** | **0.017\*** | **0.224** | **0.015\*** | **0.300** |
| **Gender** | **1591.5 † †** | **0.005\*\*** | **1055.0 † †** | **0.101** | **1055.0 † †** |
| **Social Environmental Barriers** | **0.257** | **0.008\*** | **-0.412** | **< 0.001\*\*** | **-0.488** |
| **Education (years)** | **-0.016** | **0.877** | **0.116** | **0.234** | **0.136** |
| **Marital Status** | **909.5 † †** | **0.087** | **1460.0 † †** | **0.823** | **1632.0 † †** |
| **Post-Onset months** | **-0.039** | **0.682** | **-0.093** | **0.314** | **0.003** |
| **Mobility** | **0.230** | **0.016\*** | **0.057** | **0.540** | **0.198** |
| **Depression** | **179.5 † †** | **0.350** | **180.0 † †** | **0.821** | **170.5 † †** |
| **Apathy** | **-0.039** | **0.718** | **-0.046** | **0.652** | **-0.135** |
| **Non-Verbal Intelligence** | **0.120** | **0.214** | **0.01** | **0.850** | **0.018** |
| **Social participation** | **NA** | **NA** | **0.037** | **0.688** | **0.171** |

Note: Gender and marital status were examined by Mann-Whitney U test († †: U value), and Spearman correlation assessed the other variables. \*\*p<0.01, \*p<0.05. NA; not applicable

**Table 4.** 4A. Multiple Logistic Regression Analyses on Employment Status

| Variables | $\beta$ | Standard Error | P-value | Odds ratio | 95% confidence interval for $\beta$ |
| --- | --- | --- | --- | --- | --- |
| **Aphasia Severity** | 1.301 | 0.621 | 0.036* | 3.672 | 1.087-12.410 |
| **Age** | -0.159 | 0.081 | 0.050 | 0.853 | 0.728-1.000 |
| **Non-Verbal Intelligence** | -0.300 | 0.199 | 0.131 | 0.741 | 0.502-1.094 |
| **Education** | 0.013 | 0.341 | 0.969 | 1.013 | 0.519-1.977 |
| **Apathy** | -18.673 | 7120.521 | 0.998 | 0.000 | 0.000 |

| Variables | $\beta$ | Standard Error | P-value | 95% confidence interval for $\beta$ |
| --- | --- | --- | --- | --- |
| **Mobility** | 1.423 | .472 | 0.003* | 0.489-2.360 |
| **Gender** | 1.687 | 0.545 | 0.003* | 0.607-2.768 |
| **Age** | -0.035 | 0.017 | 0.048* | -0.069-0.000 |
| **Marital status** | 0.480 | 0.509 | 0.348 | -0.530-1.490 |
| **Environmental** | 0.000 | 0.011 | 0.980 | -0.021-0.021 |

**4C. Multiple linear regression analyses performed on quality of life (SAQOL total score).**
| Variables | $\beta$ | Standard Error | P-value | 95% confidence interval for $\beta$ |
| --- | --- | --- | --- | --- |
| **Environmental barriers** | -0.013 | 0.002 | <0.001* | -0.017- -0.009 |
| **Social participation** | 0.048 | 0.021 | 0.024* | 0.006-0.090 |
| **Aphasia severity** | 0.043 | 0.021 | 0.041* | 0.02-0.084 |
| **Age** | 0.005 | 0.004 | 0.156 | -0.002-0.012 |
| **Mobility** | 0.061 | 0.088 | 0.492 | -0.114-0.236 |

**4D. Multiple linear regression analyses performed on quality of life (SAQOL Communication score).**
| Variables | $\beta$ | Standard Error | P-value | 95% confidence interval for $\beta$ |
| --- | --- | --- | --- | --- |
| **Environmental barriers** | -0.013 | 0.004 | <0.001* | -0.210- -0.006 |
| **Aphasia severity** | 0.127 | 0.034 | <0.001* | 0.061-0.194 |
| **Age** | 0.005 | 0.006 | 0.370 | -0.007-0.017 |

R<sup>2</sup> = 0.279, Adjusted R<sup>2</sup> = 0.258

### Employment Status

Employment was maintained by only 13 out of 56 participants (23%). Of these 13 participants, 11 (87%) were full-time employees, while 2 (13%) had part-time jobs. The participants primarily worked in roles such as deliveries, cleaning, simple clerical tasks like numerical data entry, and piecework. In group comparisons, the explanatory variables that had a significant negative effect on employment status were aphasia severity (p=0.004), older age (p=0.033), and apathy levels (p=0.042). The explanatory variables that did not reach statistical significance but had p-values less than 0.1 were education levels (p=0.055) and non-linguistic cognitive function (p=0.056). The small number of individuals with depression makes it statistically unavailable. Figure 1 illustrates the relation between employment status and aphasia severity, which had the strongest relation among these variables. Individuals without employment (left) exhibited more severe aphasia compared to those with employment (right). These five explanatory variables were included in the multiple logistic regression model, which clarified that only aphasia severity had a significant negative effect on employment status (p=0.036). Our logistic regression model showed a Cox-Snell R-squared of 0.443 and a Nagelkerke R-squared of 0.679, indicating that the model explains 44.3% to 67.9% of the variance in the dependent variable. The Hosmer-Lemeshow test was non-significant (p=0.732), suggesting a good fit between observed and predicted values.

**Figure 1.**
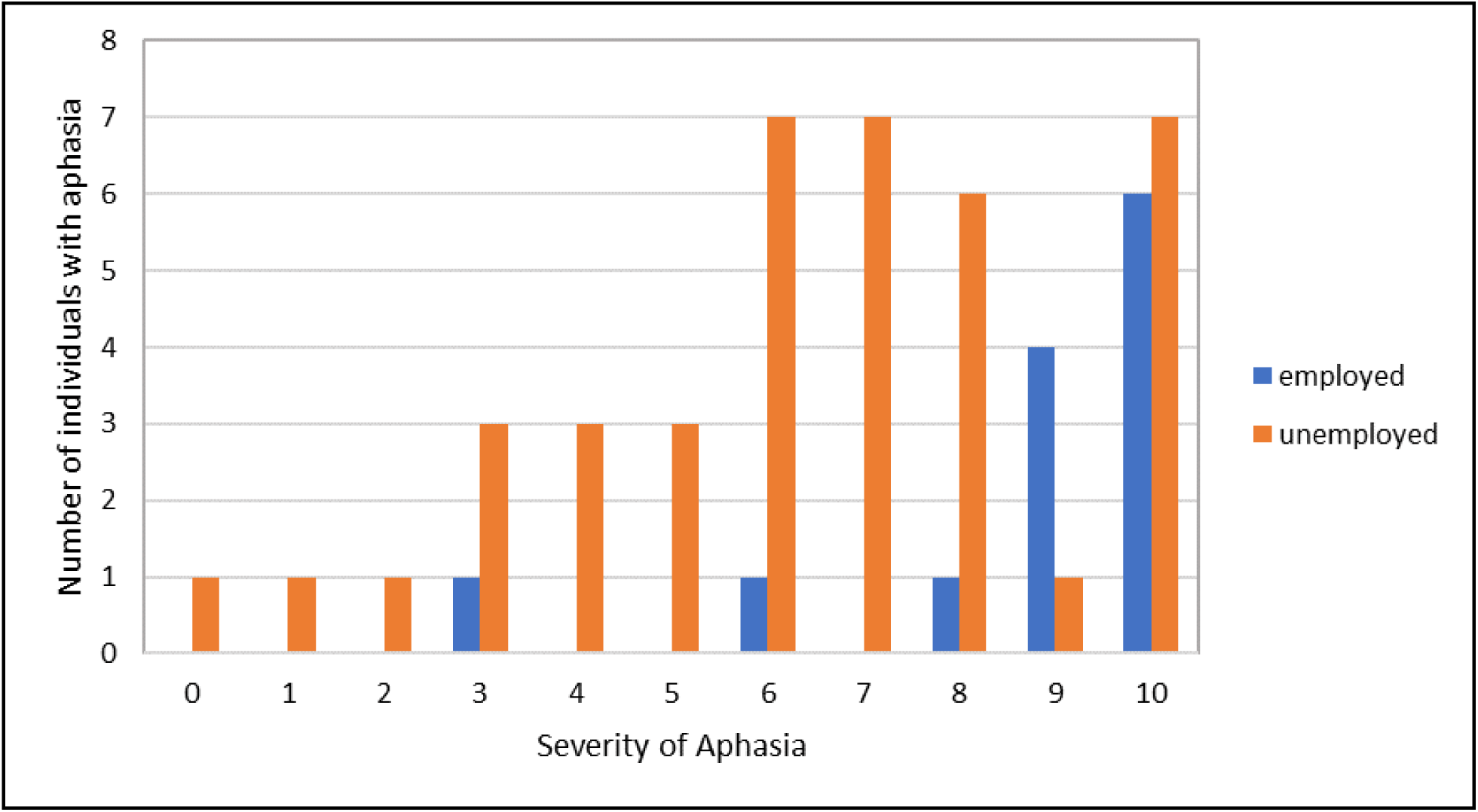
Employment Status by Aphasia Severity: Number of Employed and Unemployed Individuals with Aphasia. The blue bar represents number of employed individuals with aphasia while the orange bar represents number of those who were unemployed.

### Social Participation Levels

The average score for social participation levels in individuals in this cohort was 5.0 ± 2.0, which is similar to that reported in Lee’s study for individuals with aphasia^16^ (5.7 ± 2.0) and is below the normative data for healthy individuals in their 60s in Japan (8.4 ± 2.2) ^40^. In the simple linear regression model and Mann-Whitney U test, the variables that had a significant negative effect on social participation levels were male gender (p=0.005), high levels of social environmental barriers (p=0.008), impaired mobility (p=0.016) and older age (p=0.017). Although marital status did not reach statistical significance (p=0.087), it was included as an explanatory variable in the multiple linear regression model. The results showed that impaired mobility (p=0.003), male gender (p=0.003) and older age (p=0.048) had a significant negative impact on social participation levels. This model had an adjusted R-squared of 0.164, indicating that it explains 16.4% of the variance in the dependent variable (F=5.198, p<0.001). Regarding the relation between employment status and social participation levels, employed individuals with aphasia were not significantly different from unemployed individuals in the degree of social participation (p=0.494).

### Quality of Life

In the simple linear regression model and Mann-Whitney U test, the variables that had a significant negative effect on quality of life were younger age (p<0.001), high levels of social environmental barriers (p<0.001), aphasia severity (p=0.031), and impaired mobility (p=0.032). Low levels of social participation tended to be associated with a low quality of life (p=0.065). Regarding the relation between employment status and quality of life, there was no significant difference in quality of life between employed and unemployed individuals with aphasia (p=0.889). The five variables mentioned above, with significance and tendency, were included in the multiple linear regression model. This model showed that social environmental barriers (p<0.001), low levels of social participation (p=0.024), and aphasia severity (p=0.041) had a significant negative impact on quality of life. This model had an adjusted R-squared of 0.389, indicating that it explains 38.9% of the variance in the dependent variable (F=14.541, p<0.001).

In the sub-assessment of the communication domain in the SAQOL assessment, similar findings were observed: In the simple linear regression model and Mann-Whitney U test, the variables that had a significant negative effect on communication domain in quality of life were aphasia severity (p<0.001), high levels of social environmental barriers (p<0.001), and younger age (p=0.015). These variables were included in the multiple linear regression model, which showed that aphasia severity (p<0.001) and high levels of social environmental barriers (p<0.001) had a significant negative impact on the communication domain in quality of life. This model had an adjusted R-squared of 0.258, indicating that it explains 25.8% of the variance in the dependent variable. (F=13.421, p<0.001).

## Discussion

Our study found that aphasia severity had significantly negative impact on employment status and quality of life, particularly in the communication domain, in individuals with chronic aphasia. Additionally, social environmental barriers also negatively affected quality of life, as well as the communication domain in quality of life. Regarding social participation, low mobility, male gender, and older age were identified as negative factors. Employment status was not related to social participation levels nor quality of life.

It might seem natural to assume that aphasia severity negatively impacts employment status, but our study is the first comprehensive one to confirm this. We also found that employment status may be independent of social participation levels and quality of life. Our findings on the proportion of employment status among individuals with aphasia align with those from previous reports^1, 13, 14^. For instance, Hirali et al. ^1^ found that only 5 of 83 (6%) individuals with aphasia, who were more than 13 months post-onset and 56% of whom were of working age, were engaged in part-time jobs, volunteer work, or were students. Additionally, Naess et al. ^13^ observed that 4 out of 20 (20%) individuals with chronic aphasia who were of working age were employed. Fotiadou et al. ^14^ noted that 2 out of 10 (20%) individuals with chronic aphasia, who were between 31 to 69 years old, were employed. In our cohort, 13 out of 56 participants (23%) who were more than 18 months post-onset and of working age were employed. Despite differences in cohort characteristics, detailed work roles, regions, and time periods, these studies indicate severe employment challenges among individuals with chronic aphasia, with more than three-fourths of individuals with chronic aphasia being unemployed. The lack of an association between employment status and social participation levels suggests that these two factors may be independent and should be considered separately. This might indicate that even individuals with aphasia who are employed may not necessarily have personal social participation, such as having a confidant, visiting friends or relatives, or engaging in leisure activities with others. Alternatively, those with personal social participation may still struggle with work engagement. Additionally, the absence of an association between employment status and quality of life indicates that even when individuals with chronic aphasia are employed, they may not necessarily have a better quality of life. Rehabilitation staff, social workers, job coaches, and family members providing linguistic rehabilitation or occupational support might want to consider this when caring for individuals with chronic aphasia.

Regarding social participation, our findings were almost in line with those in Dalemans et al. ^15^. In both studies, poor mobility, male gender, and older age had a significantly negative impact on social participation among individuals with chronic aphasia. However, Dalemans et al. ^15^ found that aphasia severity was associated with poor social participation, whereas in our cohort, aphasia severity did not correlate with social participation levels. It is easy to understand that poor mobility is an important determinant of the level of social participation. Men might be more isolated than women even in the general population, as the social participation levels in the Community Integration Questionnaire are lower in men than in women in the general population, according to a study for the standardization of this questionnaire in Japan^40^. It is generally accepted that as people age their social connection weaken and consequently socially isolated. The reason why aphasia severity did not correlate with social participation in our study is unclear. Our study suggests that even mild aphasia can negatively affect social participation, such as visiting friends or relatives, engaging in leisure activities with others, and having a confidant. It is possible that personal factors such as mobility, gender and age had much stronger impact on social participation rather than aphasia severity. Further research should be conducted to clarify the inconsistent results.

Although poor mobility had a significantly negative impact on social participation, it did not correlate with employment status. This might be due to the small number of participants in the employment status analysis (N=56). Another potential reason could be the requirement for reasonable accommodation for people with disabilities in companies. This mandate requires both public and private entities to avoid unfair discriminatory treatment and to provide reasonable accommodations to remove social barriers for persons with disabilities in Japan (https://www.japaneselawtranslation.go.jp/en/laws/view/3052/en). However, this requirement does not apply to personal social participation activities such as shopping, leisure activities, or having friends or a confidant, which may explain the observed association between poor mobility and limited social participation among individuals with aphasia, but not between poor mobility and employment status.

Poor quality of life was correlated with aphasia severity, social environmental barriers, and low levels of social participation. Most of these findings are consistent with those in previous studies^1,16,19,20^. Aphasia severity also had a negative effect on quality of life among individuals with aphasia in the studies by Bullier et al. ^19^ and Filipska-Blejder et al. ^20^. The negative influence of social environmental barriers on quality of life was also demonstrated in the study by Hirali et al. ^1^. Lower level of social participation was also associated with poor quality of life in the study by Lee ^16^. Contrary to past studies, factors such as depression, education levels, and mobility did not correlate with quality of life in our study. This difference might partly be attributable to methodological aspects of our study, which will be discussed in the limitations section.

Our research underscores the substantial impact of aphasia severity on occupational and quality of life outcomes. Based on our findings, we highly recommend implementing treatments during acute phases with careful consideration of aphasia prognosis, intensive linguistic rehabilitation, and occupational support to enhance employment status and quality of life among individuals with chronic aphasia.

### Limitations

Our study has several limitations when generalizing our findings. First, we did not use a depression scale but relied on a dichotomy of the presence or absence of antidepressant medication, which limits our ability to detect mild depression. While this method effectively detects moderate to severe depression, it may overlook milder cases. Second, we did not include fatigue as an explanatory variable, despite its inclusion in some relevant studies. This decision was influenced by the inclusion of fatigue levels in the SAQOL-39, a primary outcome measure, which introduces circularity into our study. Furthermore, none of the participants in our study were employed under disability schemes, which may represent a unique employment status between being employed and unemployed. Future studies could provide insights into the underlying factors affecting employment under disability schemes among individuals with aphasia.

## Conclusions

This study found that aphasia severity negatively impacted employment status and quality of life among stroke survivors with aphasia. Employment status was not related to social participation levels nor quality of life. Our findings shed light on potential treatment options during acute phases of stroke, linguistic rehabilitation, and occupational support for individuals with chronic aphasia.

## Data Availability

All data referred to in the manuscript are available upon reasonable request from the corresponding author.

